# Beyond Guidelines: Stakeholder’s Experiences and implementation of a new model for antenatal care in Kavre district, Nepal

**DOI:** 10.1101/2025.08.03.25332922

**Authors:** KC Samita, Rajani Shakya, Abha Shrestha, Abha Shrestha, Emma Radovich

## Abstract

**Background:** In 2016, WHO updated its antenatal care (ANC) guidelines, increasing the minimum number of ANC visits from four to eight and recommending practices for those visits. The new ANC model spoke to the idea of more active connection between pregnant women and to enhance the quality and experience of ANC. Though the new ANC model was adopted in Nepal in 2020, little research has examined how the guidelines have been implemented. This study aims to explore the perspectives of pregnant women, ANC providers and local policymakers on the rollout of the new 8+ visit ANC model and identify implementation gaps between policy and the experience on the ground.

**Methods:** We conducted a convergent parallel mixed-method study involving semi-structured facility observations of ANC processes over the course of 2-3 days and key informant interviews with pregnant women, healthcare providers and local policymakers. We purposively selected six primary care level health facilities providing ANC in three municipalities in Kavre district, Nepal and conducted eight interviews with pregnant women, seven interviews with providers and three interviews with local policymakers. The interviews were audio recorded and transcribed verbatim; facility observations were documented via checklists and in field notes prepared after each observation.

**Results:** The new ANC model was perceived primarily as an increase in number of visits for the purpose of documentation. While orientations were offered on the new ANC schedule, providers reported having no orientation on content of ANC visits. There was low awareness among pregnant women about the changes in the ANC model, but women noticed more ANC appointments compared to their previous pregnancies. Pregnant women highlighted difficulty with access to health facilities, including the time and financial constraints, long waiting periods in the referred sites and physiological challenges as major challenges in adhering to increased visits.

**Conclusions:** The emphasis on quantity over quality may undermine the intended benefits of the new ANC model. There is little understanding of how the visits should be used to deliver all the recommended content of ANC. Materials to enhance understanding of intended benefits of the new ANC model would help ANC providers in health facilities to improve ANC practices. It is essential to prioritize person-centered health care during pregnancy to ensure a positive pregnancy experience. It is essential to have efforts from all stakeholders for successful implementation of the new ANC model.

**Funding source:** The study was funded by an unrestricted donation to LSHTM by Reckitt. The funder had no role in the design of the study.

**Conflict of Interest:** The study team declares no conflict of interest.

## Introduction

Antenatal care (ANC) is a crucial aspect of maternal healthcare, providing essential care and support to women during pregnancy, which provides a platform for timely interventions to prevent complications by providing health education, disease prevention, early detection and treatment, and birth preparedness.

Until 2016, the World Health Organization (WHO) had recommended a Focused Antenatal Care (FANC) model that included recommended interventions delivered over at least four ANC visits during the pregnancy. This recommendation was based on the evidence at the time suggesting that similar maternal and fetal outcomes were achieved with minimum of four targeted ANC visits compared to models with higher number of visits (1). The WHO updated its ANC guidelines in 2016 to recommend care and interventions to promote a positive experience and outcome of pregnancy and increased the minimum number of visits from four to eight during pregnancy (2). This new 8+ ANC model highlighted that women’s interactions with ANC providers should be opportunities for comprehensive care which would include medical interventions and monitoring, emotional support as well as timely information throughout the pregnancy. Hence, the WHO used the term “contact” instead of “visit” in the new ANC model, implying the idea of more active connection between women and ANC providers (2).

This new ANC model integrates clinical interventions and monitoring along with an equal emphasis on timely information, and psychosocial and emotional support by care providers (3). The changes are informed by evidence which suggests more frequent ANC visits, particularly at later stages of pregnancy, can improve the detection and management of complications such as hypertensive disorders which remains as leading cause of maternal mortality (4). Nepal adopted the new 8+ ANC model in 2022 and recommended the interventions outlined in the WHO guideline as minimum, acknowledging that these increased ANC contacts from four to eight would result in timely detection of health problems (5).

Nepal has made a significant progress in ANC attendance with the help of various health policies and programs. The overall rate of at least 4 ANC visits has increased significantly from 53% in 2011to 81% in 2022 as highlighted by Nepal Demographic Health Survey (NDHS) 2021(6). Despite the high coverage of 4+ ANC visits, few women reported 8+ ANC visits during three years of period which was just prior to the rollout of 8+ ANC model (6). The Nepal Safe Motherhood and Newborn Health Road Map 2030 emphasized that coverage of essential maternal and newborn health services has increased but quality of care remains inadequate, and the roadmap also reported that women are likely to bypass local health facilities and prefer those facilities with more health services (7). The key issues deterring the quality of ANC included poor service readiness, lengthy waiting times, and inadequate communication from providers and challenges linked with the provider’s workload, availability of training and skills (7).

While the recommended timing of ANC visits is provided, the WHO and Nepal National Medical Standard for Maternal and Newborn care guidelines did not define the content of each of those contacts. A facilitation material called “ANC to PNC continuum of care guideline” was developed on 2022 in Nepal which aided by providing detailed information regarding timing of contact and contents on history taking, examinations, interventions, health promotion activities and actions to be carried out on each visit (8).

There is limited accessible evidence on how the new WHO recommended 8+ ANC contact model is being implemented in health facilities in Nepal. In contrast, other studies have examined implementation of the new 8+ ANC model in other settings as in Mozambique and Ethiopia, finding substantial gaps in content and challenges in delivery. These included service delivery challenges-limited availability of skilled staff, poor service quality, lack of routine audits and limited awareness of guidelines, and other barriers such as low level of women’s empowerment, insufficient financial support, poor knowledge of ANC visits and inaccessibility of health facilities continued to hinder the utilization of services (9)(10). To our knowledge, this is the first study to incorporate views from policymakers, providers and pregnant women on implementation of the new 8+ ANC model.

This study aims to provide insights into the rollout of the new 8+ ANC model from the perspective of pregnant women, ANC providers and local policy makers in Nepal. This will aid in identifying implementation gaps between policy and the experience on the ground.

## Methodology

### Study design

This study was conducted in the context of an exploratory mixed method research project examining antenatal urine screening practices. This study involved face-to-face interviews with pregnant women, ANC providers and local policymakers and semi-structured facility observations at the selected health facilities.

### Study Setting

ANC services in Nepal are accessible from all public health facilities including community level primary healthcare outreach clinics (11). The study was conducted in three municipalities (two urban and one rural) of Kavre district, Bagmati province. All municipalities have started implementing the new 8+ ANC model, where the schedule of visits differs from WHO recommended schedule as show in Table 1. (5)

**Table 1:** Comparing the recommended timing of ANC visits in WHO and Nepal guidelines.

| Timing of contacts | WHO FANC model | WHO 8+ Recommended schedule | Nepal 4 ANC model | Nepal new ANC schedule |
| --- | --- | --- | --- | --- |
| Contact 1 | 8-12 weeks | Up to 12 weeks | 12-16 weeks | Up to 12 weeks |
| Contact 2 | 24-26 weeks | 20 weeks | 20-24 weeks | Up to 16 weeks |
| Contact 3 | 32 weeks | 26 weeks | 28-32 weeks | 20-24 weeks |
| Contact 4 | 36-38 weeks | 30 weeks | 36-40 weeks | 28 weeks |
| Contact 5 | - | 34 weeks |  | 32 weeks |
| Contact 6 | - | 36 weeks |  | 34 weeks |
| Contact 7 | - | 38 weeks |  | 36 weeks |
| Contact 8 | - | 40 weeks |  | 38-40 weeks |

### Sampling and Study participants

We purposively selected government health facilities in the municipalities based on ANC case flow, and range of services offered by health facilities such as lab and birthing center services. Local policymakers were also consulted about selecting the study sites. Two health facilities were selected in each municipality (a total of six facilities).

Similarly, we purposively selected the study participants. We interviewed pregnant women visiting the health facilities for ANC, providers who were particularly involved in providing ANC services and the local policymakers to gain deeper insights into the findings from pregnant women and ANC PROVIDERs from selected health facilities.

### Data collection

Data collections were conducted by two researchers: SK-a sociologist and RS-a clinically trained researcher. We conducted semi-structured observations and face-to-face interviews in each participating health facility. We developed a semi-structured observation checklist to gain an in-depth understanding of the interaction between pregnant women and providers during ANC visits, and to collect information on facility physical infrastructure, including water and sanitation infrastructure. Between two to three days were spent at all six health facilities, prioritizing ANC days for observation to further aid the interview process. This approach of observation led to capturing real-time interactions and ANC environments to understand the contextual factors shaping ANC provision within selected health facilities. The semi-structured observation checklist was pretested in the ANC clinic of a tertiary level hospital in the district and adapted accordingly. Observations were conducted in the health facilities prior to interviews with providers.

Interviews were conducted based on topic guides prepared for each type of study participant. The topic guide for pregnant women and ANC Providers was as pretested in the ANC clinic of a tertiary-level hospital in the district and adapted accordingly. The researchers also iteratively adapted the interview topic guides based on field notes collected during the observations. Along with this, reflection meetings with the research team were conducted after completing data collection in each facility, identifying further areas for investigation and probing in subsequent interviews.

The data collection was done from January to June 2024.

### Data analysis

All the interviews were audio recorded and transcribed verbatim. The interviews were analyzed in Nepali language to preserve linguistic nuances that could be lost in the translation. Initial codes were developed inductively by SK and were reviewed and discussed with the research team. A subset of the interviews was translated into English and double coded by SK and ER. Iterative process involved coding, developing and refining codes and themes. Relevant sections of the transcripts were translated into English for the write up of findings.

Interviews of different types of participants were coded separately and then the themes were compared and interrogated to generate a richer picture of 8+ ANC model implementation. These themes were later also interrogated with observation reports to triangulate findings.

A stakeholder meeting of policymakers and representatives from participating health facilities was conducted in July 2024 to share preliminary findings from the analysis. The insights from stakeholders were used to reflect on and validate initial interpretations and to refine the final version of results.

We drew from the Theory of Acceptability (12) during the data analysis, which offered a conceptual lens to interpreting various stakeholder’s perception on implementation of new 8+ ANC model. However, the framework did not adequately align with our data hence we adopted an inductive thematic approach without anchoring our findings to the theory.

### Ethical consideration

Written informed consent was taken before the interviews from each participant whereas written informed consent with facility in charge was taken prior to the facility observations. Ethical approval for the study was obtained from the Nepal Health Research Council (Reg No: 635/2023), Kathmandu University Institutional Review Committee (Approval No: 85/24) and London School of Hygiene and Tropical Medicine (LSHTM Ethics Ref: 29823). The voluntary participation of all the study participants was ensured throughout the study process.

## Results

The study took place in six health facilities offering ANC services in three municipalities (Table 1). In total, eight interviews with pregnant women, seven interviews with ANC PROVIDERs and three interviews with local policymakers were conducted.

**Table 1:** Study setting and number of study participants.

| Type of Health Facilities | Number of Health Facilities | Total Pregnant women Interviewed | Total ANC PROVIDERs interviewed |
| --- | --- | --- | --- |
| Health Post<sup>a</sup> | 4 | 4 | 4 |
| Primary Health Care Center | 1 | 3 | 1 |
| Municipal Hospital | 1 | 1 | 2 |
| **Total** | **6** | **8** | **7** |

<sup>a</sup> All but one of the health posts did not have a birthing unit.

All the included health facilities offered childbirth services except for one health post. All seven providers interviewed were female and were engaged primarily in providing ANC services to pregnant women visiting the health facilities. The eight pregnant women participating in study were between four to nine months pregnant and ranged in age from 19-36 years old. The educational backgrounds of pregnant women varied from primary level to bachelor’s degree, including two participants who were not able to read and write. The participants involved pregnant women who were pregnant for the first, second and third times during their lifetime. Among the women, three were pregnant for the first time, three were pregnant for the second time and two for the third time.

Three overarching themes were derived during the process of data analysis. The first theme centered around how the policy was understood by the ANC providers and local policymakers as reflecting the idea that the new ANC model was primarily about an increase in number of visits for documentation. The second theme centered on how inadequate orientation to the new 8+ ANC model hindered the practices of providers on providing quality ANC care. The third theme centered around the barriers pregnant women experience while attending multiple ANC visits.

### Theme 1: Understanding policy as primarily an increase in numbers of documentation of ANC visits

ANC providers appeared to understand the new ANC model as a mandate to document the number of ANC contacts in the provided Safe Motherhood register. ANC providers emphasized mostly about the importance of documenting ANC visits in register to follow the schedule of ANC visit to provide incentives to women attending scheduled ANC visits. ANC incentives included certain amount of money that are given by providers for pregnant women upon completion of scheduled ANC visits.

One of the ANC providers stated it as:

> *“There is our schedule of 8 visits, and we are calling according to that schedule because when they (Pregnant women) come, if they do not visit as per the protocol then they will not get that benefit (ANC incentive) given by the government. Like, if their visit is as per the protocol then they would get 800 rupees after delivery as nutrition cost and we remind them to come as per the schedule, if they came as per schedule they would get that benefit.” -ANC Provider1*

Similarly, the other ANC Provider stated:

> *“If they (pregnant women) come as per the protocol, then we could find out their complications which might be there, and they would also get to have iron and calcium in regular basis and would not miss it and also the target of government would be achieved, and they would get the benefit(ANC incentive) given by government if they did not miss any one of the visits.” -ANC Provider4*

Though the ANC providers were aware that the increased contact would lead to screening of complications, it was found that ANC providers were not aware of the reasons behind increasing visits also included a need to develop monitoring indicators that focus on the content of ANC as part of quality care. It was found that providers did not note any significant changes in the two different ANC models.

One of the ANC providers highlighted the similarities and differences between 8 ANCs and 4 ANCs as:

> *“The whole checkup is the same, I did not see anything new on 8 ANCs like this should be done specially in this month. There might be and we might not have been trained on it, that is in its own place. We used to take weight before and measure it now as well, anomaly was done earlier as well and is done now too, now the VDRL (Venereal Disease Research Laboratory) lab tests were done earlier as well and now too. There is nothing like, this has been added specially in 8 visits.” -ANC Provider7*

Most ANC Providers also expressed that they had been facing issues on recording the ANC visit and having problems with the new register developed for 8 ANC. One of the ANC providers stated it as:

> *“Before this came, we heard like now Government of Nepal is bringing 8 visit and after that we heard that it was implemented and even now also we are uninformed on regular recording and doing as much as we can…difficulty is like, first meeting with them remains as first meeting right, and that person would have come in 13 weeks, one pregnant mother would come in 13 weeks but in the place to record its given as before 12 weeks and then 16 weeks, then it would be like whether to do it in 12 weeks or 16 weeks, the person is of 13 weeks who have already crossed 12 weeks so cannot take to 12 weeks and also cannot take to 16 weeks, it’s difficult. Now the focal person above us is also confused on recording these cases, it’s not that we did not ask, we asked.”*-ANC Provider7

On the other hand, local policymakers having the role of supervising and monitoring the providers were also largely unfamiliar recommended guidelines of 8 ANC. One of the local policymakers also stated it as:

> *“No, there are no such differences, the tests which are done during the 4 visits, the same tests and counseling are done in 8 visits. The times of visit increased and in cases of weight increment they get to see the same activities 4 times more.”-Local Policy Maker 2*

Similarly, one of the policymakers raised a question: “Is it necessary to train for all things?” upon enquiring about if there is provision of any ANC training packages to providers highlighting a significant concern among providers who on the other hand find training requirement as a major barrier to delivering quality care.

### Theme 2: Positive views upon change of ANC model but little orientation

Providers in the health facilities stated that the increased number of ANC visits had improved their relationship with pregnant women, indicating that they believed they were providing good care. They also spoke of the potential benefits of more visits, reflecting the positive implications of the change in policy. One of the providers stated it as:

> *What I felt is, when they needed to come many times then there would be a little more intimacy (connection) and our service seeker would express their problems more openly and I felt that belongingness more. The more times they need to come, they would feel like this is their own institution, so, they need to visit it and tell us about problems, there is like… they tell their problems more eagerly. I feel like this might have happened and there is an increase in intimacy with them. I feel like it has been a little easier for me. -ANC Provider5*

However, it was observed that the recommended guidelines for the content of these visits were not being followed adequately since providers implementing the ANC model were not familiar with the recommended content of care in the guideline but were aware about the shift in schedule.

One of the ANM primarily involved in providing ANC stated:

> *“The training given at first was 4 times and on 8 times checkup, what should be examined and why it became 8 times for this once orientation was given to nursing staff in the municipality. The other sister went at that time, she came and told us the things said there but let’s say the training was not given more carefully saying about what should be examined on 8 times to all nursing staff providing ANC in the institution”. – ANC Provider1*

It was observed that despite the inconsistency in care across the health facilities, some of the ANC providers were offering personalized attention to the pregnant women. When pregnant women reported any health issues, these providers extended consultation time and offered information accordingly to the women. However, in the absence of expressed concerns, counselling remained limited.

Additionally, none of the providers were aware of the availability of facilitating materials indicating all recommended services on 8 ANCs, highlighting the significant gap in knowledge and adherence to best ANC practice. Providers implementing the new ANC model had different experiences as some found it challenging whereas some did not. They claimed that they had not been well oriented towards implementing the new ANC model.

One of the ANC providers narrated about gaps in orientation before implementation as:

> *“Ahuh, we have not known. It’s just said that 8 visits need to be made. Now we are calling regularly according to the time (schedule) and doing the things that we have been doing earlier but there has not been any specialized orientation to us on what should be done on which visit, nothing has happened to us.”-ANC Provider3*

Providers also spoke of the need for more training/orientation on policy changes and stated the need for materials that would guide their practices while providing services.

One of the ANC providers stated it as,

> *“We had talked about it (availability of information materials) as well, if there is availability of flip chart indicating what should be done on what time, sometime there will be much patient flow and we do not get chance to work freely, sometimes we may have confusion so if we had a book or a flipchart then I feel like it would be easier. No, we have not been provided with that.” – ANC provider4*

Nevertheless, the motivated workforce or the intrinsic motivation towards work on the other hand also do have a profound impact on policy implementation. Notably, one of the ANC providers who on her own initiative created information materials outlining the recommended guidelines and was consistently providing services in alignment. The provider claimed the lack of proper orientation from higher authorities, however, contributed on her own to enhancing the quality of care. She stated it as:

> *“It was said that the checkup should be done 8 times, the weeks were told briefly but it was not said in detail… We don’t have a protocol; we took it out on our own saying that it’s necessary. We also need to copy from others, I used to go to family planning (implying one of the other organization) and used to look there and I came back by capturing a photo and said them that it should be kept like this, after I said so, as there are junior sisters here who have more knowledge than me, I told them and they took it out accordingly and we pasted it.”-ANC Provider 6*

Amidst concerns about inadequate orientation impacting providers’ ability to deliver quality service, a local policymaker emphasized that the orientation/training as referred by service providers before implementation of new 8+ ANC model would be orientation which lacks resource materials. The policymaker highlighted that proper training would include participant’s handouts.

The local policy maker stated it as:

> *If it’s the training, then there would be manuals and there would be books for participants. Now, in this orientation, one may get but they may get the PowerPoints of the presentation and other things, but I don’t think there would be separate exact books or protocols (In orientation).* – Local Policy Maker 4

However, the study team had located an implementation guide called “ANC to PNC Continuum of Care Guideline” online that was not available in any of the facilities during the time of observation. This was discussed in the stakeholder’s meeting where local policymakers were largely unaware of the document but responded positively to the resource materials. The policymakers expressed a desire to use the materials as soon as possible and advocated for its proper distribution.

One of the local policymakers in an interview expressed this unawareness and narrated it as:

> *One of the reasons for arising this sort of gap(inconsistency in services of ANC) is, all of us became irresponsible, all of us mean those who developed the guideline, it should have been reached out to concerned places and assured whether that(guideline) reached there or not, for example, did they made it reach to my district or not and did I give it to local level or not? and even if you (Researcher) find it out after searching then the person providing the ANC check would have also found it if searched, so they (Providers) are also irresponsible. Meaning that all of us became irresponsible. – Local policy maker 4*

### Theme 3: High efforts but low returns

Pregnant women were largely unfamiliar with the new model and most of them explained that they had been visiting the health facilities for ANC appointments as per the instruction of providers. Some women noticed and appreciated an increase in the number of contacts compared to previous pregnancy experiences. A pregnant woman who was seeking her ANC in last month of the pregnancy compared the new model to her previous experience:

> *“Now, I feel good. When I gave birth to my daughter, it was about 8,9 years ago. Now it’s said that one needs to check up to 8,9 months and till before having the baby. When given birth to my daughter I just came 2-3 times, video Xray and nothing else was done. Now it’s said that what things need to be done. I am doing whatever they say.” - Pregnant Woman3*

Though pregnant women believed that ANC contacts are beneficial for them, observation done in ANC settings revealed that not all facilities offer satisfactory ANC services. One of the facilities offered minimal consultation time, as brief as three minutes of interaction between the provider and women. Similarly, it was also noted in the observation that women rarely received recommended counselling during their ANC visits including the recommended investigations as routine urine screening.

Pregnant women highlighted the challenges as distant health facility from the place of residence, difficulty with access to transportation, difficulty to manage household chores, financial constraints when referred to facility that costs money, long waiting periods in the referred sites (tertiary level hospitals) and physiological challenges as pains in leg and back as major concerns for them adhering to multiple ANC visits. A pregnant woman narrated:

> *“Now I don’t have any person in my house to look after me, I have one daughter and I have been raising a few goats at home and I feel a little like, how many times to walk…like that. I have been called 4 times during my daughter’s turn and now it’s not like before also.”-Pregnant Woman3*

Additionally, one of the ANC providers from health facility belonging to rural areas narrated the challenges of pregnant women on behalf of her experiences with ANC client as:

> *“How I feel is, Bichara (helpless people), they do have challenges, why not? because they need to come by walking from far distance and its much difficult for them when it comes at last last (late months of pregnancy), there is no facility of vehicle for those Bichara, like when coming from that far, there is uphill as well as downhill ways and they need to come and go by hanging that big stomach which is difficult. Now, when we ask them to come after two weeks, they say that it’s much more difficult and you call every time. And we need to send them by counselling that as we have protocol it’s difficult, but you need to come during this time as the difficulties (risks) might arise. They say things like how many times you call and how many times to come. They feel bored and gets irritated.”-ANC Provider6*

However, suboptimal quality of ANC services which does not include adequate content of care during each visit makes the efforts of women less beneficial when they attend health facilities despite multiple challenges. It was found during the observation in ANC setting that the pregnant women attending the ANC visits were not provided with all content of care as mentioned in the national guideline of providing ANC. Routine examinations, such as urine testing in each visit, and counselling were missing from most ANC consultations observed.

## Discussion

The study explored the perspective of policymakers, providers and pregnant women on the roll out of the 8+ ANC model in Kavre district, Nepal. The findings suggested a critical gap between the understanding of model’s changes to both the purpose and frequency of ANC contacts among all stakeholders, along with gaps in implementation of recommended content of care. These gaps contribute significantly to suboptimal ANC quality despite increased efforts of pregnant women to complete additional ANC visits.

Our findings suggested that the providers and local policymakers had limited understanding of rationale behind change in the ANC model, seeing the model as solely about an increase in number of ANC visits. While the WHO recommended ANC visit schedule emphasized frequent monitoring particularly on the final weeks of pregnancy to address risks like hypertension and other complications which emerges closer to the childbirth(4), the recommended visit schedule from Government of Nepal deviated by including higher number of contacts earlier in the pregnancy, particularly in second trimester (2)(5). This shift suggests that Nepal’s guideline might have missed the critical objective of frequent monitoring in final weeks of pregnancy, raising the important concern about whether this ANC schedule effectively addresses the key health issues as designed by WHO.

We found that providers and stakeholders had limited understanding about the content of care recommendations for each visit. This echoes findings from other low and middle income countries which showed that coverage of care components during ANC visits is often lower than high coverage of ANC attendance suggesting that quality of ANC interventions has not been given as much attention as achieving minimum numbers of visits (13)(14) (15). A study in Ethiopia found similar challenges faced by providers and local policymakers during roll out of 8+ ANC model (10), highlighting barriers such as: unavailability of guideline, inadequate pre-service clinical supervision, poor quality of in-service training, lack of regular mentorship and supervision and lack of monitoring and evaluation as on adhering to ANC guidelines despite the willingness of provider to provide care. Similar to our findings, the study in Ethiopia emphasizes that the implementation of the new 8+ANC model requires comprehensive roll out strategies to support adoption of the full guidance for content of care in the 8+ ANC model (10). Consequently, implementation without the corresponding orientation for providers and support for monitoring not just the visit schedule but also the delivery of recommended ANC intervention the successful implementation of the model and its intended benefits.

Our findings align with Downe and colleagues’ (2016) review which highlighted that women pursue ANC with goals for a positive pregnancy and motherhood experience and this includes ensuring a healthy pregnancy, maintaining physical and sociocultural normality and receiving emotional and psychosocial support (3). In our study, women were highly motivated to attend prescribed ANC visits, driven by strong desire to protect their health and that of baby. They went to great effort to attend ANC visits more frequently under the 8+ ANC model. Our findings emphasize the need to increase accessibility of ANC services but also responsive, respectful and person-centered ANC services in facilities which acknowledge women’s efforts in seeking services. A study in Malawi presented their findings under seven constructs of the Theoretical Framework of Acceptability and found that the new ANC model was acceptable among women, though they were unaware about existence of the new model, but noted the burden on women associated with increased ANC visits (cite Malawi study). Specifically, long distance of healthcare facilities and lack of transportation were major obstacles discussed by women (12), echoing findings from our study.

### Strengths and Limitations of the study

The study drew on multiple approaches to qualitative data collection including semi-structured observations and interviews with a variety of stakeholders. Interviews conducted during the facility observation visits allowed comparison of collected information through the process of capturing real-time interaction between providers and pregnant women. The study validated its findings with stakeholders later to reflect and refine interpretations.

Despite the strengths, the study had a few limitations. The study was conducted in the context of an exploratory study about ANC WASH (Water, Sanitation and Hygiene) infrastructure and provision of urine testing in pregnancy. Issues of implementation of the 8+ ANC policy emerged out of early data collection, so initial interviews did not focus on this topic. Topic guides were iteratively refined during data collection in response to emerging findings as the research team developed greater understanding of implementation challenges.

The data collection was conducted at health posts, which remains the first point of contact for health services in the community. However, the study was conducted in a smaller number of facilities of few municipalities and did not include higher level health facilities, so we were unable to examine implementation across all levels of health facilities involved in ANC.

The researchers made efforts to build rapport with pregnant women during interviews, however, the process of interviews was quite challenging. Only some pregnant women were willing to participate in interviews as many of them had time constraints and other commitments. The pregnant women who participated also had limited time and expressed worry about keeping their visitors waiting during the interviews, which may have inhibited responses. Additionally, the combined power dynamics in terms of educational background of interviewer and participant, particularly the rural women with varying educational level and social positioning might have impacted the richness and openness of discussion. Furthermore, the facility-based data collection limits understanding of pregnant women who are not able to overcome access barriers.

### Conclusions and recommendations

The study integrated findings from facility observations and interviews with stakeholders in Kavre district, Nepal on the roll out of the 8+ ANC model. Implementation of the new model primarily focused on increasing the number of visits, where limited knowledge of providers on the recommended content of each visit led to the observed suboptimal quality of ANC contacts. It is crucial for providers to make each ANC contact meaningful especially given the access barriers pregnant women face. Improved training and orientation for providers could support greater awareness of the purpose of the new ANC model and greater adherence to guideline-recommended care. Implementing tangible tools such as facilitation materials, flip charts, posters, pamphlets is essential to assist providers in adhering to ANC protocols. However, these efforts must be complemented by supportive monitoring, supervision and a conducive environment. Addressing the gap in content of care is essential to realize the intended benefits of the new 8+ANC model which contributes to improved maternal and perinatal outcomes to ensure a positive pregnancy experience.

Lastly, the introduction of increased ANC visits in the 8+ ANC model calls for improved maternal care while its successful implementation depends on addressing systemic barriers, provider’s preparedness and adequate awareness among all stakeholders. Therefore, our finding highlights the importance of aligning policy goals with frontline realities to ensure positive pregnancy experience for all pregnant women as advocated by updated guidelines.

## Data Availability

Data that supports the findings of this study can be made available by corresponding author upon reasonable request.

## Acknowledgements

This work was funded by an unrestricted donation to LSHTM by Reckitt. The funder had no role in the design of the study.

